# Case fatality risk of the SARS-CoV-2 variant of concern B.1.1.7 in England

**DOI:** 10.1101/2021.03.04.21252528

**Authors:** Daniel J Grint, Kevin Wing, Elizabeth Williamson, Helen I McDonald, Krishnan Bhaskaran, David Evans, Stephen JW Evans, Alex J Walker, George Hickman, Emily Nightingale, Anna Schultze, Christopher T Rentsch, Chris Bates, Jonathan Cockburn, Helen J Curtis, Caroline E Morton, Sebastian Bacon, Simon Davy, Angel YS Wong, Amir Mehrkar, Laurie Tomlinson, Ian J Douglas, Rohini Mathur, Paula Blomquist, Brian MacKenna, Peter Ingelsby, Richard Croker, John Parry, Frank Hester, Sam Harper, Nicholas J DeVito, Will Hulme, John Tazare, Ben Goldacre, Liam Smeeth, Rosalind M Eggo

**Author notes:** **Corresponding author:** Daniel Grint.

## Abstract

The B.1.1.7 variant of concern (VOC) is increasing in prevalence across Europe. Accurate estimation of disease severity associated with this VOC is critical for pandemic planning. We found increased risk of death for VOC compared with non-VOC cases in England (HR: 1.67 (95% CI: 1.34 - 2.09; P<.0001)). Absolute risk of death by 28-days increased with age and comorbidities. VOC has potential to spread faster with higher mortality than the pandemic to date.

## Introduction

The SARS-CoV-2 variant of concern B.1.1.7 (VOC) was first identified in Kent, UK in autumn 2020. Early analysis suggests it is more transmissible than previously circulating forms (non-VOC).^1^ It is now the dominant strain throughout the UK and is increasing in prevalence across Europe.^2^ Early reports of increased mortality have not included data on individuals’ comorbidities, and this information is needed to facilitate pandemic planning.

In specific types of PCR assays for SARS-CoV-2, missingness in one spike protein gene target occurs with this VOC. Spike gene target failure (SGTF) is therefore a proxy for VOC identification, with greater than 95% sensitivity during the period 16^th^ November – 11^th^ January.^3^

Working on behalf of NHS England, we estimate the risk of death following confirmation of SARS-CoV-2 infection in England, comparing infection with VOC to non-VOC, after accounting for demographic factors and comorbidities. The code and configuration of our analysis is available online (github.com/opensafely/sgtf-cfr-research).

### Study population

Data were drawn from the OpenSAFELY electronic health records (EHR) secure research platform, covering 40% of England’s population registered with a general practitioner (GP) (see Supplement 1.). We used linked GP, SARS-CoV-2 testing, vaccination and mortality data (Supplement Table S1).

Vaccinations against and diagnoses of SARS-CoV-2 prior to the study period were exclusion criteria. SGTF status was known for 184,786/441,161 (42%) people with confirmed SARS-CoV-2 infection between 16^th^ November 2020 and 11^th^ January 2021 (91,775 VOC; 93,011 non-VOC) (Supplement Table S4). Full details of the design and analysis are available in the protocol (Supplement 9.). A total of 867 (419 VOC; 448 non-VOC) all-cause deaths occurred prior to 5^th^ February 2021. The exposure groups were similar demographically (Table 1). The VOC group was younger with a lower proportion of older cases (80+: 0.9% VOC vs. 1.6% non-VOC cases), with fewer comorbidities (2+ comorbidities: 2.9% vs. 3.8%). Non-VOC cases were more frequent in the first four weeks of the study period, while VOC cases predominated thereafter. Consequently, median follow-up time was shorter among the VOC group (36 days (interquartile range (IQR): 30-45)) than the non-VOC group (57 days (40-72)).

**Table 1.** Summary demographic and clinical characteristics of the study population. A full table including all factors adjusted for is given in Supplementary Table S3.

|  | **Total**<br>**N (%)** | **non-VOC cases**<br>**N (%)** | **VOC cases**<br>**N (%)** |
| --- | --- | --- | --- |
| **Total population** | 184,786 | 91,775 | 93,011 |
| **Deaths** | 867 (0.5) | 448 (0.5) | 419 (0.5) |
| **Time to death** |  |  |  |
| Median (IQR) | 13.0 (9.0-21.0) | 13.0 (8.0-22.0) | 14.0 (9.0-21.0) |
| **Follow-up time** |  |  |  |
| Median (IQR) | 43.0 (33.0-60.0) | 57.0 (40.0-72.0) | 36.0 (30.0-45.0) |
| **Epidemiological week of diagnosis** |  |  |  |
| 16 Nov-22 Nov | 21,976 (11.9) | 20,854 (22.7) | 1,122 (1.2) |
| 23 Nov-29 Nov | 14,755 (8.0) | 13,432 (14.6) | 1,323 (1.4) |
| 30 Nov-06 Dec | 14,286 (7.7) | 11,576 (12.6) | 2,710 (2.9) |
| 07 Dec-13 Dec | 18,137 (9.8) | 11,703 (12.8) | 6,434 (6.9) |
| 14 Dec-20 Dec | 19,963 (10.8) | 9,043 (9.9) | 10,920 (11.7) |
| 21 Dec-27 Dec | 24,422 (13.2) | 8,246 (9.0) | 16,176 (17.4) |
| 28 Dec-03 Jan | 34,527 (18.7) | 9,477 (10.3) | 25,050 (26.9) |
| 04 Jan-11 Jan | 36,720 (19.9) | 7,444 (8.1) | 29,276 (31.5) |
| **Sex** |  |  |  |
| Female | 98,099 (53.1) | 49,468 (53.9) | 48,631 (52.3) |
| Male | 86,687 (46.9) | 42,307 (46.1) | 44,380 (47.7) |
| **Age group** |  |  |  |
| 18-<30 | 36,969 (20.0) | 17,302 (18.9) | 19,667 (21.1) |
| 30-<40 | 34,298 (18.6) | 16,782 (18.3) | 17,516 (18.8) |
| 40-<50 | 32,783 (17.7) | 15,904 (17.3) | 16,879 (18.1) |
| 50-<60 | 30,484 (16.5) | 15,261 (16.6) | 15,223 (16.4) |
| 60-<70 | 14,818 (8.0) | 7,587 (8.3) | 7,231 (7.8) |
| 70-<80 | 5,860 (3.2) | 3,116 (3.4) | 2,744 (3.0) |
| 80+ | 2,346 (1.3) | 1,513 (1.6) | 833 (0.9) |
| **Categorical number of comorbidities<sup>a</sup>** |  |  |  |
| 0 | 158,017 (85.5) | 77,538 (84.5) | 80,479 (86.5) |
| 1 | 20,606 (11.2) | 10,768 (11.7) | 9,838 (10.6) |
| 2+ | 6,163 (3.3) | 3,469 (3.8) | 2,694 (2.9) |
| **Index of multiple deprivation** |  |  |  |
| 1 least deprived | 36,560 (19.8) | 15,973 (17.4) | 20,587 (22.1) |
| 2 | 34,767 (18.8) | 16,000 (17.4) | 18,767 (20.2) |
| 3 | 35,181 (19.0) | 16,192 (17.6) | 18,989 (20.4) |
| 4 | 38,603 (20.9) | 19,479 (21.2) | 19,124 (20.6) |
| 5 most deprived | 39,675 (21.5) | 24,131 (26.3) | 15,544 (16.7) |

<sup>a</sup>Comorbidities as defined in Supplementary Table S2.

### Relative hazard of death

We calculated the relative hazard of death for VOC compared to non-VOC cases using a Cox proportional hazards regression model stratified by region (upper tier local authority area (UTLA)). Follow-up was censored on the 5^th^ February 2021 or seven days prior to receipt of a SARS-CoV-2 vaccine, whichever came first. VOC was consistently associated with an increased hazard of death. In fully adjusted analysis accounting for demographics and comorbidities, hazards were two-thirds higher in the VOC group (hazard ratio (HR): 1.67 (95% confidence interval (CI): 1.34 – 2.09; P <0.0001)) (Figure 1). Increased hazards were consistent across all pre-specified subgroup analyses including epidemiological week, age group, categorical number of comorbidities, ethnicity, and deprivation index quintile. Increased hazards were also consistent across all sensitivity analyses; in analysis restricted to people diagnosed with SARS-CoV-2 a minimum of 28-days prior to the censoring date the hazard ratio was 1.71 (95% CI: 1.36 – 2.15; P <0.0001).

**Figure 1.**
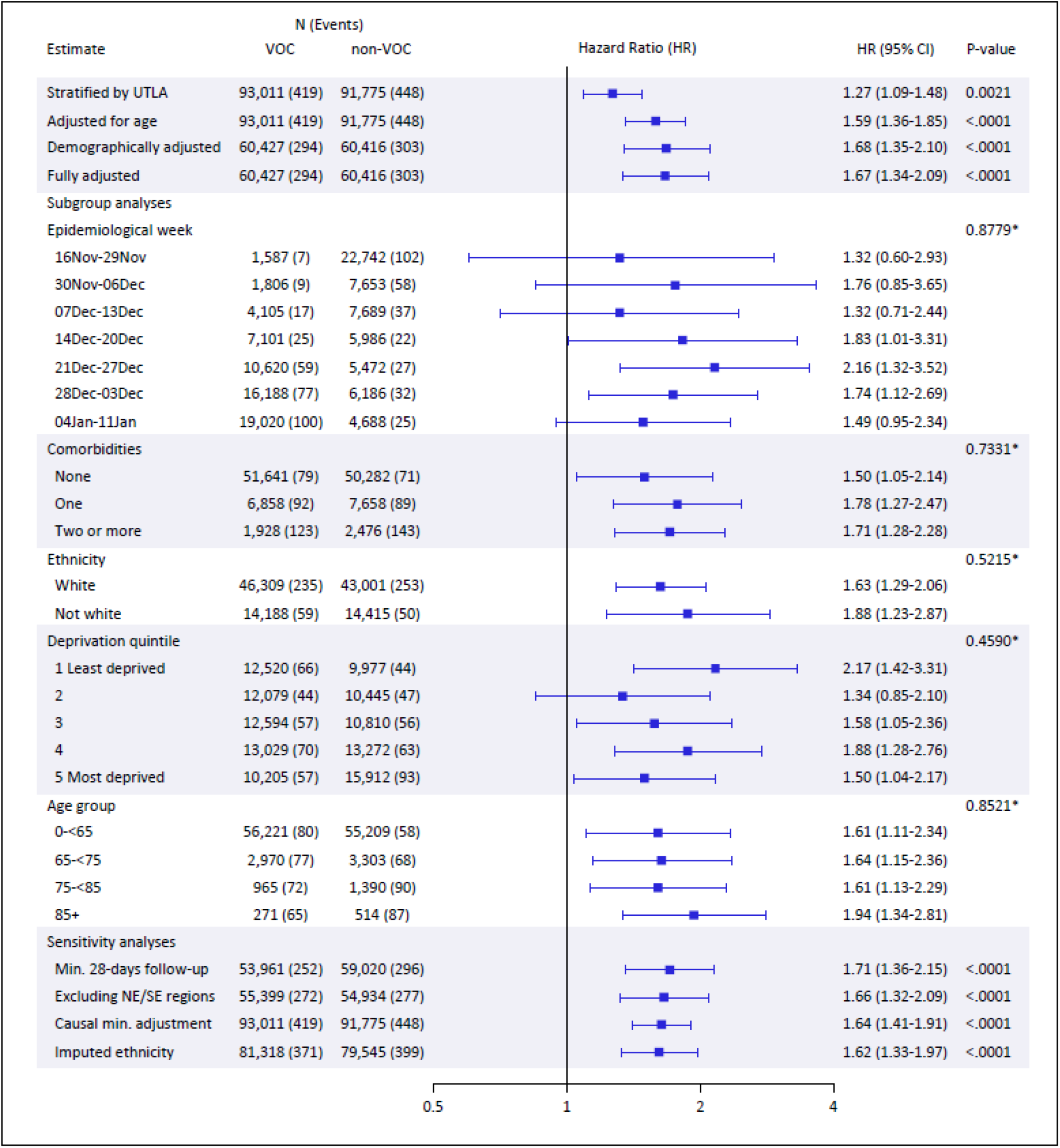
Hazard ratios for VOC vs. non-VOC from Cox proportional hazards regression stratified by Upper Tier Local Authority (UTLA). All subgroup analyses were performed on the fully adjusted model. *Likelihood ratio test for interaction between exposure group (VOC, non-VOC) and subgroup All models are stratified on region by UTLA. Demographically adjusted model includes adjustment for: age, sex, IMD, ethnicity, household size, rural urban classification, epidemiological week, and care home status. The fully adjusted model includes adjustment for: age, sex, IMD, ethnicity, smoking status, obesity, household size, rural urban classification, comorbidities, epidemiological week, and care home status. There was no evidence of non-proportional hazards in this model (global test of Schoenfeld residuals, P=0.19). The first sensitivity analysis is restricted to people with a minimum of 28-days from testing positive for SARS-CoV-2 to the follow-up censor. The South East and North East NHS England regions are excluded from the second sensitivity analysis (details Supplement 7.). The causal minimum adjustment set includes adjustment for: age, care home status, comorbidities, deprivation index, and smoking status. Missing ethnicity data was imputed for the final listed sensitivity analysis.

### Absolute risk of death by 28-days

We found a higher absolute risk of death by 28-days post SARS-CoV-2 positive test by age, sex, and presence of comorbidities in VOC (Table 2). Risk of death was estimated by the marginal means of a fully adjusted logistic regression model with the outcome of death within 28-days; restricted to people diagnosed with SARS-CoV-2 a minimum of 28-days prior to the censoring date. Consistent with the Cox model, VOC was associated with increased odds of death in this model (adjusted odds ratio: 1.73 (95% CI: 1.34 - 2.23; P-value <.0001), vs. non-VOC). The risk of death was low for those aged under 65 in the absence of comorbidities, though higher for males than females (28-day risk of death for those aged <65 with no comorbidities: (VOC: Males 0.14%; Females: 0.07%); (non-VOC: Males: 0.09%; Females: 0.05%)). The risk of death was consistently higher for males and increased with age and the presence of comorbidities. The highest risk was seen among those aged 85 and above with two or more comorbidities (28-day risk of death for those aged 85+ with two or more comorbidities: VOC: (Males 24.3%; Females: 14.7%); (non-VOC: Males: 16.7%; Females: 9.7%)). The excess risk of death by 28-days for VOC compared to non-VOC is shown in Figure 2.

**Table 2.** Absolute risk of death by 28-days. Absolute risk is calculated from the marginal means of a fully adjusted logistic regression model with outcome death by 28-days, restricted to the population with a minimum of 28-days from testing positive for SARS-CoV-2 to the follow-up censor. The fully adjusted model includes adjustment for: age, sex, IMD, ethnicity, smoking status, obesity, household size, NHS England region, rural/urban classification, comorbidities, epidemiological week, and care home status.

|  | **non-VOC**<br>**% (95% CI)** | **VOC**<br>**% (95% CI)** |
| --- | --- | --- |
| **No Comorbidities** |  |  |
| **Female: 0-<65** | 0.05 (0.03-0.06) | 0.07 (0.06-0.09) |
| **65-<75** | 0.45 (0.30-0.59) | 0.72 (0.50-0.95) |
| **75-<85** | 1.08 (0.71-1.45) | 1.73 (1.15-2.31) |
| **85+** | 2.36 (1.47-3.25) | 3.75 (2.34-5.16) |
| **Male: 0-<65** | 0.09 (0.07-0.11) | 0.14 (0.11-0.17) |
| **65-<75** | 0.85 (0.59-1.12) | 1.37 (0.96-1.77) |
| **75-<85** | 2.03 (1.35-2.71) | 3.24 (2.19-4.30) |
| **85+** | 4.38 (2.72-6.03) | 6.87 (4.33-9.42) |
| **1 Comorbidity** |  |  |
| **Female: 0-<65** | 0.11 (0.08-0.15) | 0.18 (0.13-0.24) |
| **65-<75** | 1.09 (0.78-1.41) | 1.75 (1.25-2.25) |
| **75-<85** | 2.60 (1.84-3.35) | 4.13 (2.94-5.32) |
| **85+** | 5.54 (3.77-7.31) | 8.64 (5.91-11.38) |
| **Male: 0-<65** | 0.22 (0.15-0.28) | 0.35 (0.25-0.45) |
| **65-<75** | 2.06 (1.51-2.62) | 3.29 (2.44-4.14) |
| **75-<85** | 4.81 (3.48-6.14) | 7.54 (5.52-9.55) |
| **85+** | 9.94 (6.87-13.01) | 15.10 (10.63-19.58) |
| **2+ Comorbidities** |  |  |
| **Female: 0-<65** | 0.21 (0.14-0.28) | 0.34 (0.22-0.45) |
| **65-<75** | 1.99 (1.41-2.57) | 3.18 (2.27-4.09) |
| **75-<85** | 4.66 (3.45-5.87) | 7.31 (5.42-9.20) |
| **85+** | 9.65 (7.01-12.29) | 14.68 (10.73-18.63) |
| **Male: 0-<65** | 0.40 (0.27-0.52) | 0.64 (0.44-0.84) |
| **65-<75** | 3.72 (2.74-4.69) | 5.87 (4.38-7.35) |
| **75-<85** | 8.44 (6.44-10.44) | 12.93 (9.99-15.87) |
| **85+** | 16.65 (12.42-20.88) | 24.34 (18.55-30.13) |

**Figure 2.**
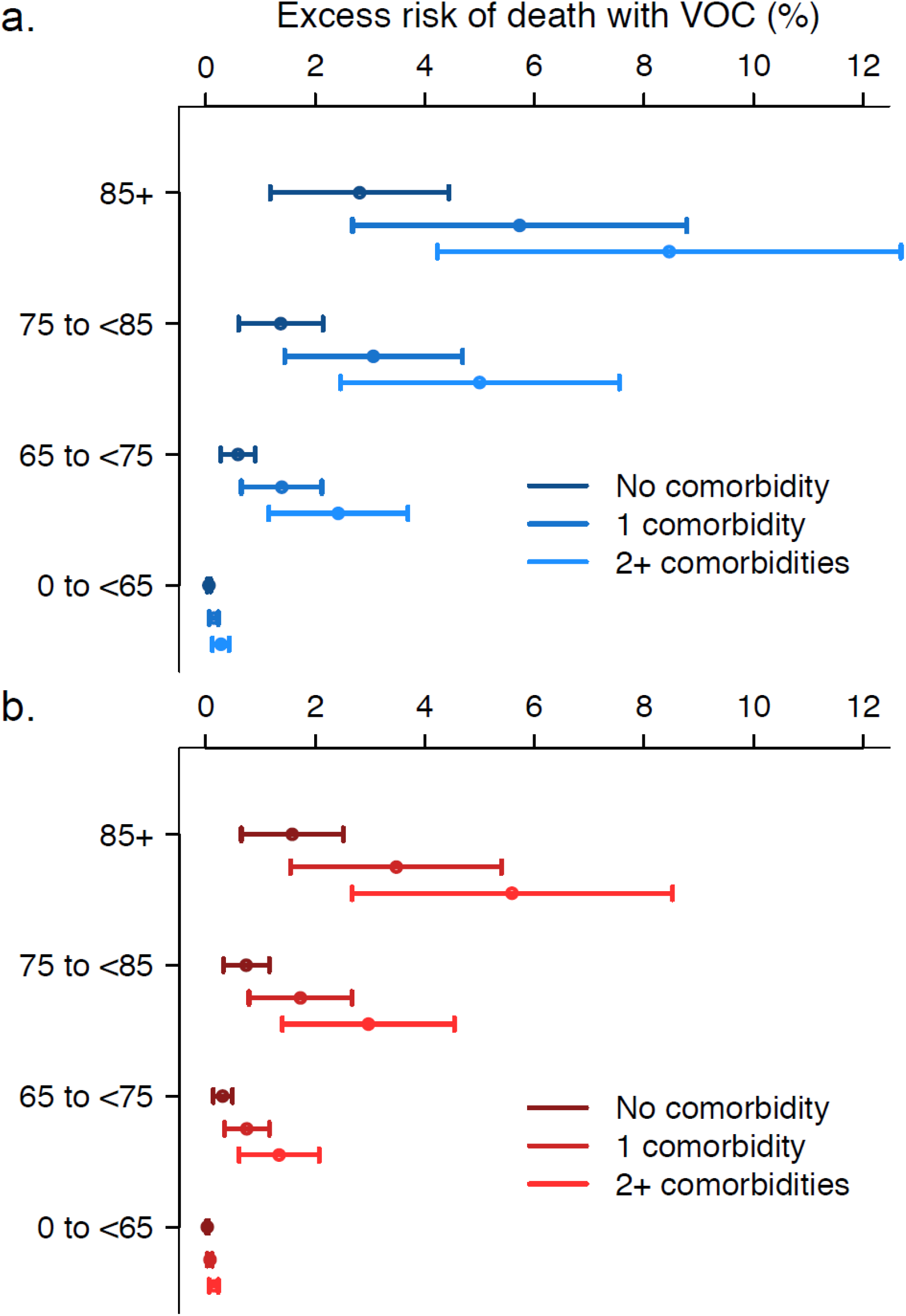
Excess risk of death by 28-days for VOC compared to non-VOC. The risk difference of death by 28-days for VOC compared to non-VOC, with 95% confidence interval, in a: Males; b: Females

## Discussion

The SARS-CoV-2 B.1.1.7 VOC has been the subject of intense research since its emergence. Increased transmissibility means it is now the most common variant in the UK, a trend confirmed here. We find this VOC was associated with two-thirds higher case fatality than the previously circulating virus in this unvaccinated population. For every three deaths in a population with the previously circulating virus we would expect five deaths in a similar population with VOC. Other studies have assessed the relative mortality of the VOC with similar conclusions,^4-6^ however, our results are the first to include detailed information on the presence of comorbidities. Interestingly, the effects of age and comorbidities appear to be collinear as adjustment for comorbidities did not alter the findings after adjustment for age. As prevalence of many comorbidities is associated with age this finding appears plausible.^7^ The consistency of the effect for all calendar time periods shows that the increase in mortality due to VOC could not be explained by other secular changes in mortality, such as hospitals exceeding capacity.

The absolute risks of death by 28-days demonstrate the increasing risks with age and presence of comorbidities, while males have consistently higher risk of death than females. However, age and comorbidity risk factors associated with poor non-VOC outcomes appear to be similar to those with this VOC. Therefore, prioritisation for vaccination and shielding can remain the same.

In the UK, all-cause death by 28-days post confirmation of SARS-CoV-2 infection is the standard definition of SARS-CoV-2 mortality,^8^ so we used death from any cause as the primary outcome. In sensitivity analysis restricted to people diagnosed with SARS-CoV-2 a minimum of 28-days prior to the censoring date, and logistic regression with deaths censored beyond 28-days the results were consistent.

This VOC is now prevalent across Europe and is likely to become the most frequent variant following the pattern seen in the UK.^2^ Policy makers and planners need to account for higher mortality of this VOC.

Crucially, emerging data suggest that the currently approved vaccines for SARS-CoV-2 are effective against the B.1.1.7 VOC.^9^ This study highlights the importance of robust national vaccination programmes and infection control measures to contain the SARS-CoV-2 pandemic. Unmitigated spread of the B.1.1.7 VOC has the potential to be both faster and more deadly than the pandemic to date.

## Data Availability

The data are stored in a secure platform and cannot be shared directly. Please see opensafely.org for access information.

## Ethical approval

This study was approved by the Health Research Authority (REC reference 20/LO/0651) and by the LSHTM Ethics Board (reference 21863).

## Acknowledgements

We are grateful for the support received from the TPP Technical Operations team and for generous assistance from the information governance and database teams at NHS England / NHSX. We would also like to thank PHE for making SARS-CoV-2 testing data available, along with Daniel West and Jamie Lopez-Bernal for processing and sharing the SGTF data.

## Author contributions

DG, RE, and KW led the study. RE conceived the study. DG, RE, KW, KB, CR, and LS drafted the study protocol. DG, KW, RE, EW, HMcD, KB, DE, SE, AJW, EN, AS, CR, HC, CM, AYSW, RM, PB, WH, and JT contributed to data preparation and variable definitions. DE, GH, CB, JC, HC, CM, SB, SD, AM, LT, ID, BMcK, PI, RC, JP, FH, SH, NDeV, WH, BG, and LS contributed to building the analytical platform. DG, KW, RE, KB, SE, and LS contributed to study design. DG performed the statistical analysis. All authors contributed to manuscript preparation and refinement.

## References

1. Davies NG, Abbott S, Barnard RC, et al. Estimated transmissibility and impact of SARS-CoV-2 lineage B.1.1.7 in England. Science 2021: eabg3055.

2. European Centre for Disease Prevention and Control (ECDC): SARS-CoV-2 - increased circulation of variants of concern and vaccine rollout in the EU/EEA,14th update. https://www.ecdc.europa.eu/sites/default/files/documents/RRA-covid-19-14th-update-15-feb-2021.pdf, 2021.

3. Public Health England (PHE): Investigation of novel SARS-CoV-2 variant. Variant of concern 20201/01. 2020. https://assets.publishing.service.gov.uk/government/uploads/system/uploads/attachment_data/file/959438/Technical_Briefing_VOC_SH_NJL2_SH2.pdf (accessed 25/2/2021 2021).

4. NERVTAG. NERVTAG paper on COVID-19 variant of concern B.1.1.7: Paper from the New and Emerging Respiratory Virus Threats Advisory Group (NERVTAG) on new coronavirus (COVID-19) variant B.1.1.7. https://www.gov.uk/government/publications/nervtag-paper-on-covid-19-variant-of-concern-b117.

5. Iacobucci G. Covid-19: New UK variant may be linked to increased death rate, early data indicate. British Medical Journal Publishing Group; 2021.

6. Davies NG, Jarvis CI, Edmunds WJ, Jewell NP, Diaz-Ordaz K, Keogh RH. Increased mortality in community-tested cases of SARS-CoV-2 lineage B.1.1.7. medRxiv 2021: 2021.02.01.21250959.

7. Walker JL, Grint DJ, Strongman H, et al. UK prevalence of underlying conditions which increase the risk of severe COVID-19 disease: a point prevalence study using electronic health records. medRxiv 2020: 2020.08.24.20179192.

8. Department of Health and Social Care: New UK-wide methodology agreed to record COVID-19 deaths. https://www.gov.uk/government/news/new-uk-wide-methodology-agreed-to-record-covid-19-deaths: gov.uk; 2020.

9. Public Health England (PHE): PHE monitoring of the effectiveness of COVID-19 vaccination. 2021. https://www.gov.uk/government/publications/phe-monitoring-of-the-effectiveness-of-covid-19-vaccination (accessed 25/2/2021 2021).

