## Supplementary material for "Case fatality risk of the SARS-CoV-2 variant of concern B.1.1.7 in England"

#### Table of Contents


1. Further information on OpenSAFELY 2
2. Information governance and ethics 2
3. Details on data sources 2
4. Definition of comorbidities 3
5. Table S3 Complete demographic and clinical characteristics 5
6. SGTF test results during study period 7
7. SGTF positivity in TPP compared to PHE 8
8. References 9
9. Study Protocol 10

#### 1. Further information on OpenSAFELY

All data were linked, stored and analysed securely within the OpenSAFELY platform https://opensafely.org/. The dataset analysed within OpenSAFELY is based on 24 million people currently registered with GP surgeries using TPP SystmOne software. Data include pseudonymized data such as coded diagnoses, medications and physiological parameters. No free text data are included. All code is shared openly for review and re-use under MIT open license (https://github.com/opensafely/SGTF-CFR-research). Detailed pseudonymised patient data is potentially re-identifiable and therefore not shared. We rapidly delivered the OpenSAFELY data analysis platform without prior funding to deliver timely analyses on urgent research questions in the context of the global Covid-19 health emergency: now that the platform is established we are developing a formal process for external users to request access in collaboration with NHS England; details of this process will be published shortly on OpenSAFELY.org.

#### 2. Information governance and ethics

NHS England is the data controller; TPP is the data processor; and the key researchers on OpenSAFELY are acting on behalf of NHS England. This implementation of OpenSAFELY is hosted within the TPP environment which is accredited to the ISO 27001 information security standard and is NHS IG Toolkit compliant;[1,2] patient data has been pseudonymised for analysis and linkage using industry standard cryptographic hashing techniques; all pseudonymised datasets transmitted for linkage onto OpenSAFELY are encrypted; access to the platform is via a virtual private network (VPN) connection, restricted to a small group of researchers; the researchers hold contracts with NHS England and only access the platform to initiate database queries and statistical models; all database activity is logged; only aggregate statistical outputs leave the platform environment following best practice for anonymisation of results such as statistical disclosure control for low cell counts.[3] The OpenSAFELY research platform adheres to the obligations of the UK General Data Protection Regulation (GDPR) and the Data Protection Act 2018. In March 2020, the Secretary of State for Health and Social Care used powers under the UK Health Service (Control of Patient Information) Regulations 2002 (COPI) to require organisations to process confidential patient information for the purposes of protecting public health, providing healthcare services to the public and monitoring and managing the COVID-19 outbreak and incidents of exposure; this sets aside the requirement for patient consent.[4] Taken together, these provide the legal bases to link patient datasets on the OpenSAFELY platform. GP practices, from which the primary care data are obtained, are required to share relevant health information to support the public health response to the pandemic, and have been informed of the OpenSAFELY analytics platform.

This study was approved by the Health Research Authority (REC reference 20/LO/0651) and by the LSHTM Ethics Board (reference 21863).

#### 3. Details on data sources

We used several linked data sets in this analysis (Table S1). OpenSAFELY also contains linked hospitalisation and emergency department data, but those are not used in this analysis, because neither are available in near real-time unlike GP, testing, mortality and vaccination data, which are available after a short lag of 10-16 days.

| Data | Description |
|---|---|
| SARS-CoV-2 tests | Positive and negative tests of people tested under the UK's Pillar 1 and Pillar 2 testing schemes are reported.[5] Pillar 2 generally is community tests and Pillar 1 is tests in hospital (patients and health care workers). Only some of the labs used for testing in England use the 3 channel PCR for which a "failure to detect the Spike-gene target" is indicative of the VOC, therefore not all positive tests have known SGTF status. The data come from Public Health England's (PHE) Second Generation Surveillance System. |
| General Practitioner (GP) data | GP data are drawn from patients who are registered at a practice that runs the TPP SystmOne (https://www.tpp-uk.com/products/systmone). This is approximately 40% of GPs in England. Each patient encounter with a GP is coded using CTV3 codes, which fully aligns with SNOMED-CT which describe the reason for the encounter, and these codes are used to define the health history of each individual.[6] Prescribed medications are also stored in the health record. Demographic data such as age and ethnicity are collected by GPs, as are some behavioural data like whether an individual smokes. |
| Mortality date | The date of death plus codes for the cause of death are from the Office for National Statistics. We only use date of death in this study. |
| Vaccination date | The date, dose number, vaccine manufacturer and batch are entered into their health record. We only use the date of administration of the first dose in this study. |
| Index of multiple deprivation (IMD) | We use the England IMD which is matched to individuals at the postcode level. |
| Urban/Rural classification | We use 5 categories of Urban/Rural classifications which are matched to individuals at the postcode level. |

**Table S1**. Data sources used in this analysis.

#### 4. Definition of comorbidities

A patient is identified as having a comorbidity if their health record includes codes indicative of each of the conditions (Table S2).

| Condition | Codelist defining presence of comorbidity |
|---|---|
| Aplastic anaemia | https://codelists.opensafely.org/codelist/opensafely/aplastic-anaemia/ |
| Asplenia | https://codelists.opensafely.org/codelist/opensafely/asplenia/ |
| Asthma | https://codelists.opensafely.org/codelist/opensafely/asthma-diagnosis/<br>https://codelists.opensafely.org/codelist/opensafely/asthma-inhaler-salbutamol-medication/2020-04-15/<br>https://codelists.opensafely.org/codelist/opensafely/asthma-inhaler-steroid-medication/2020-04-15/<br>https://codelists.opensafely.org/codelist/opensafely/asthma-oral-prednisolone-medication/2020-04-27/ |
| Bone marrow transplant | https://codelists.opensafely.org/codelist/opensafely/bone-marrow-transplant/2020-04-15/ |
| Cancer | |

| | |
|---|---|
| Chronic cardiac disease<br>Chronic respiratory disease<br>Chronic liver disease<br>Dementia<br>Diabetes<br>Chronic kidney disease<br>GI bleed<br>HIV<br>Permanent immunosuppression<br>Temporary immunosuppression<br>Hypertension<br>Stroke<br>Inflammatory bowel disease<br>Neurological conditions<br>Psoriasis<br>Sickle cell disease<br>Smoking<br>Organ transplant | https://codelists.opensafely.org/codelist/opensafely/cancer-excluding-lung-and-haematological/2020-04-15/<br>https://codelists.opensafely.org/codelist/opensafely/chemotherapy-or-radiotherapy-updated/2020-04-15/<br>https://codelists.opensafely.org/codelist/opensafely/haematological-cancer/2020-04-15/<br>https://codelists.opensafely.org/codelist/opensafely/lung-cancer/2020-04-15/<br>https://codelists.opensafely.org/codelist/opensafely/chronic-cardiac-disease/2020-04-08/<br>https://codelists.opensafely.org/codelist/opensafely/chronic-respiratory-disease/2020-04-10/<br>https://codelists.opensafely.org/codelist/opensafely/chronic-liver-disease/2020-06-02/<br>https://codelists.opensafely.org/codelist/opensafely/dementia/2020-04-22/<br>https://codelists.opensafely.org/codelist/opensafely/diabetes/2020-04-15/<br>https://codelists.opensafely.org/codelist/opensafely/chronic-kidney-disease/2020-04-14/<br>https://codelists.opensafely.org/codelist/opensafely/gi-bleed-or-ulcer/2020-04-08/<br>https://codelists.opensafely.org/codelist/opensafely/hiv/2020-07-13/<br>https://codelists.opensafely.org/codelist/opensafely/permanent-immunosuppression/2020-06-02/<br>https://codelists.opensafely.org/codelist/opensafely/temporary-immunosuppression/2020-04-24/<br>https://codelists.opensafely.org/codelist/opensafely/hypertension/2020-04-28/<br>https://codelists.opensafely.org/codelist/opensafely/stroke-updated/2020-06-02/<br>https://codelists.opensafely.org/codelist/opensafely/inflammatory-bowel-disease/2020-04-07/<br>https://codelists.opensafely.org/codelist/opensafely/other-neurological-conditions/2020-06-02/<br>https://codelists.opensafely.org/codelist/opensafely/ra-sle-psoriasis/2020-04-14/<br>https://codelists.opensafely.org/codelist/opensafely/sickle-cell-disease/2020-04-14/<br>https://codelists.opensafely.org/codelist/opensafely/smoking-clear/2020-04-29/<br>https://codelists.opensafely.org/codelist/opensafely/smoking-unclear/2020-04-29/<br>https://codelists.opensafely.org/codelist/opensafely/solid-organ-transplantation/2020-04-10/ |

**Table S2**. Comorbidities and the codelists that determine if a patient is classified as having that comorbidity. All codelists are reviewed by clinicians.

### 5. Table S3 Complete demographic and clinical characteristics

| | Total<br>N (%) | non-VOC<br>N (%) | VOC<br>N (%) |
|---|---|---|---|
| **Total population** | 184,786 | 91,775 | 93,011 |
| **Died** | 867 (0.5) | 448 (0.5) | 419 (0.5) |
| **Time to death** | | | |
| Mean (SD) | 16.2 (11.2) | 16.8 (13.3) | 15.6 (8.3) |
| Median (IQR) | 13.0 (9.0-21.0) | 13.0 (8.0-22.0) | 14.0 (9.0-21.0) |
| **Follow-up time** | | | |
| Mean (SD) | 47.1 (17.4) | 55.8 (17.8) | 38.5 (11.8) |
| Median (IQR) | 43.0 (33.0-60.0) | 57.0 (40.0-72.0) | 36.0 (30.0-45.0) |
| **Epidemiological week of diagnosis** | | | |
| 16Nov-22Nov | 21,976 (11.9) | 20,854 (22.7) | 1,122 (1.2) |
| 23Nov-29Nov | 14,755 (8.0) | 13,432 (14.6) | 1,323 (1.4) |
| 30Nov-06Dec | 14,286 (7.7) | 11,576 (12.6) | 2,710 (2.9) |
| 07Dec-13Dec | 18,137 (9.8) | 11,703 (12.8) | 6,434 (6.9) |
| 14Dec-20Dec | 19,963 (10.8) | 9,043 (9.9) | 10,920 (11.7) |
| 21Dec-27Dec | 24,422 (13.2) | 8,246 (9.0) | 16,176 (17.4) |
| 28Dec-03Jan | 34,527 (18.7) | 9,477 (10.3) | 25,050 (26.9) |
| 04Jan-11Jan | 36,720 (19.9) | 7,444 (8.1) | 29,276 (31.5) |
| **Sex** | | | |
| Female | 98,099 (53.1) | 49,468 (53.9) | 48,631 (52.3) |
| Male | 86,687 (46.9) | 42,307 (46.1) | 44,380 (47.7) |
| **Age (years)** | | | |
| Mean (SD) | 38.2 (18.1) | 38.5 (18.5) | 37.9 (17.7) |
| Median (IQR) | 38.0 (24.0-52.0) | 38.0 (24.0-52.0) | 37.0 (24.0-51.0) |
| **Grouped age** | | | |
| 18-<30 | 36,969 (20.0) | 17,302 (18.9) | 19,667 (21.1) |
| 30-<40 | 34,298 (18.6) | 16,782 (18.3) | 17,516 (18.8) |
| 40-<50 | 32,783 (17.7) | 15,904 (17.3) | 16,879 (18.1) |
| 50-<60 | 30,484 (16.5) | 15,261 (16.6) | 15,223 (16.4) |
| 60-<70 | 14,818 (8.0) | 7,587 (8.3) | 7,231 (7.8) |
| 70-<80 | 5,860 (3.2) | 3,116 (3.4) | 2,744 (3.0) |
| 80+ | 2,346 (1.3) | 1,513 (1.6) | 833 (0.9) |
| **Ethnicity** | | | |
| White | 105,428 (57.1) | 52,687 (57.4) | 52,741 (56.7) |
| South Asian | 21,562 (11.7) | 11,880 (12.9) | 9,682 (10.4) |
| Black | 4,530 (2.5) | 1,753 (1.9) | 2,777 (3.0) |
| Mixed | 2,628 (1.4) | 1,175 (1.3) | 1,453 (1.6) |
| Other | 2,974 (1.6) | 1,351 (1.5) | 1,623 (1.7) |
| Missing | 47,664 (25.8) | 22,929 (25.0) | 24,735 (26.6) |
| **Evidence of obesity**[a] | | | |
| No record of obesity | 144,246 (78.1) | 70,584 (76.9) | 73,662 (79.2) |
| Obese I (30-34.9) | 24,536 (13.3) | 12,664 (13.8) | 11,872 (12.8) |
| Obese II (35-39.9) | 10,386 (5.6) | 5,446 (5.9) | 4,940 (5.3) |

| | | | |
|---|---|---|---|
| Obese III (40+) | 5,618 (3.0) | 3,081 (3.4) | 2,537 (2.7) |
| **Smoking status[b]** | | | |
| Never | 115,458 (62.5) | 57,727 (62.9) | 57,731 (62.1) |
| Former | 49,974 (27.0) | 24,907 (27.1) | 25,067 (27.0) |
| Current | 19,354 (10.5) | 9,141 (10.0) | 10,213 (11.0) |
| **Categorical number of comorbidities** | | | |
| No comorbidity | 158,017 (85.5) | 77,538 (84.5) | 80,479 (86.5) |
| 1 comorbidity | 20,606 (11.2) | 10,768 (11.7) | 9,838 (10.6) |
| 2+ comorbidities | 6,163 (3.3) | 3,469 (3.8) | 2,694 (2.9) |
| **Index of multiple deprivation** | | | |
| 1 least deprived | 36,560 (19.8) | 15,973 (17.4) | 20,587 (22.1) |
| 2 | 34,767 (18.8) | 16,000 (17.4) | 18,767 (20.2) |
| 3 | 35,181 (19.0) | 16,192 (17.6) | 18,989 (20.4) |
| 4 | 38,603 (20.9) | 19,479 (21.2) | 19,124 (20.6) |
| 5 most deprived | 39,675 (21.5) | 24,131 (26.3) | 15,544 (16.7) |
| **Categorical household size** | | | |
| 1-2 | 47,573 (25.7) | 24,042 (26.2) | 23,531 (25.3) |
| 3-5 | 92,701 (50.2) | 45,057 (49.1) | 47,644 (51.2) |
| 6-10 | 18,689 (10.1) | 9,469 (10.3) | 9,220 (9.9) |
| 11+ | 1,942 (1.1) | 1,007 (1.1) | 935 (1.0) |
| Missing | 23,881 (12.9) | 12,200 (13.3) | 11,681 (12.6) |
| **Care home status[c]** | | | |
| Private home | 184,312 (99.7) | 91,425 (99.6) | 92,887 (99.9) |
| Care home | 474 (0.3) | 350 (0.4) | 124 (0.1) |
| **NHS England region** | | | |
| East | 44,757 (24.2) | 9,731 (10.6) | 35,026 (37.7) |
| East Midlands | 26,530 (14.4) | 18,708 (20.4) | 7,822 (8.4) |
| London | 15,998 (8.7) | 4,503 (4.9) | 11,495 (12.4) |
| North East | 7,015 (3.8) | 5,791 (6.3) | 1,224 (1.3) |
| North West | 25,043 (13.6) | 14,743 (16.1) | 10,300 (11.1) |
| South East | 9,432 (5.1) | 2,620 (2.9) | 6,812 (7.3) |
| South West | 9,058 (4.9) | 4,893 (5.3) | 4,165 (4.5) |
| West Midlands | 15,196 (8.2) | 7,601 (8.3) | 7,595 (8.2) |
| Yorkshire and the Humber | 31,700 (17.2) | 23,140 (25.2) | 8,560 (9.2) |
| **Rural urban classification** | | | |
| Urban major conurbation | 57,872 (31.3) | 28,106 (30.6) | 29,766 (32.0) |
| Urban minor conurbation | 8,895 (4.8) | 6,811 (7.4) | 2,084 (2.2) |
| Urban city and town | 90,880 (49.2) | 43,239 (47.1) | 47,641 (51.2) |
| Rural town and fringe | 16,375 (8.9) | 8,526 (9.3) | 7,849 (8.4) |
| Rural village and dispersed | 9,102 (4.9) | 4,119 (4.5) | 4,983 (5.4) |
| Missing | 1,662 (0.9) | 974 (1.1) | 688 (0.7) |

[a]59,080 Missing values set to no evidence of obesity; [b]28,020 missing values set to never smoking status;
[c]Based on address linkage with CQC data as of 1st Feb 2020: 1,662 missing values set to private home.

#### 6. SGTF test results during study period

Positive SARS-CoV-2 test results can be assessed for SGTF if detected using the Thermo Fisher TaqPath PCR assay, which targets 3 genes including the S gene, where CT values are ≤30 for detected genes. Three Pillar 2 lighthouse laboratories report their TaqPath result to PHE's second generation surveillance system (SGSS): Alderley Park, Glasgow, and Milton Keynes laboratories. As such, available SGTF data excludes Pillar 1 tests, Pillar 2 TaqPath tests carried out by other laboratories or with high CT values, and lateral flow tests (LFT). These excluded positive results are represented by unclassified or blank values in the table below. Positive test results are classified as non-VOC if all three genes (S, ORF1ab, and N) are detected with CT values ≤30. Positive test results are classified as SGTF (VOC proxy) if the S gene is undetectable and N/ORF1ab genes are detected with CT values ≤30. LFT use has been increasing in the UK over the study period.

We are grateful for the hard work processing these data at each of the three lighthouse laboratories mentioned above.

| | **Total** | **non-VOC** | **VOC** | **Unclassified** | **Blank** |
|---|---|---|---|---|---|
| **N** | 441,161 | 91,775 | 93,011 | 207,025 | 49,350 |
| **16Nov-22Nov** | 36,394 (8.2) | 20,854 (22.7) | 1,122 (1.2) | 9,680 (4.7) | 4,738 (9.6) |
| **23Nov-29Nov** | 26,554 (6.0) | 13,432 (14.6) | 1,323 (1.4) | 7,748 (3.7) | 4,051 (8.2) |
| **30Nov-06Dec** | 26,010 (5.9) | 11,576 (12.6) | 2,710 (2.9) | 7,553 (3.6) | 4,171 (8.5) |
| **07Dec-13Dec** | 34,391 (7.8) | 11,703 (12.8) | 6,434 (6.9) | 11,233 (5.4) | 5,021 (10.2) |
| **14Dec-20Dec** | 53,838 (12.2) | 9,043 (9.9) | 10,920 (11.7) | 27,391 (13.2) | 6,484 (13.1) |
| **21Dec-27Dec** | 63,881 (14.5) | 8,246 (9.0) | 16,176 (17.4) | 33,089 (16.0) | 6,370 (12.9) |
| **28Dec-03Jan** | 97,234 (22.0) | 9,477 (10.3) | 25,050 (26.9) | 54,912 (26.5) | 7,795 (15.8) |
| **04Jan-11Jan** | 102,859 (23.3) | 7,444 (8.1) | 29,276 (31.5) | 55,419 (26.8) | 10,720 (21.7) |

**Table S4**. The number of SARS-CoV-2 positive tests in total, with stratification by epidemiological week and SGTF test result.

#### 7. SGTF positivity in TPP compared to PHE

The proportion of positive tests with SGTF in PHE data[7] and in OpenSAFELY in each epidemiological week is very similar (Figure S1). Discrepancies arise due to the geographic variation in which GP providers use TPP software. There are relatively fewer TPP SGTF positives in the NHS regions South East (especially northern Kent) and North East (especially the area around Cumbria), both of which experienced early increases in SGTF.

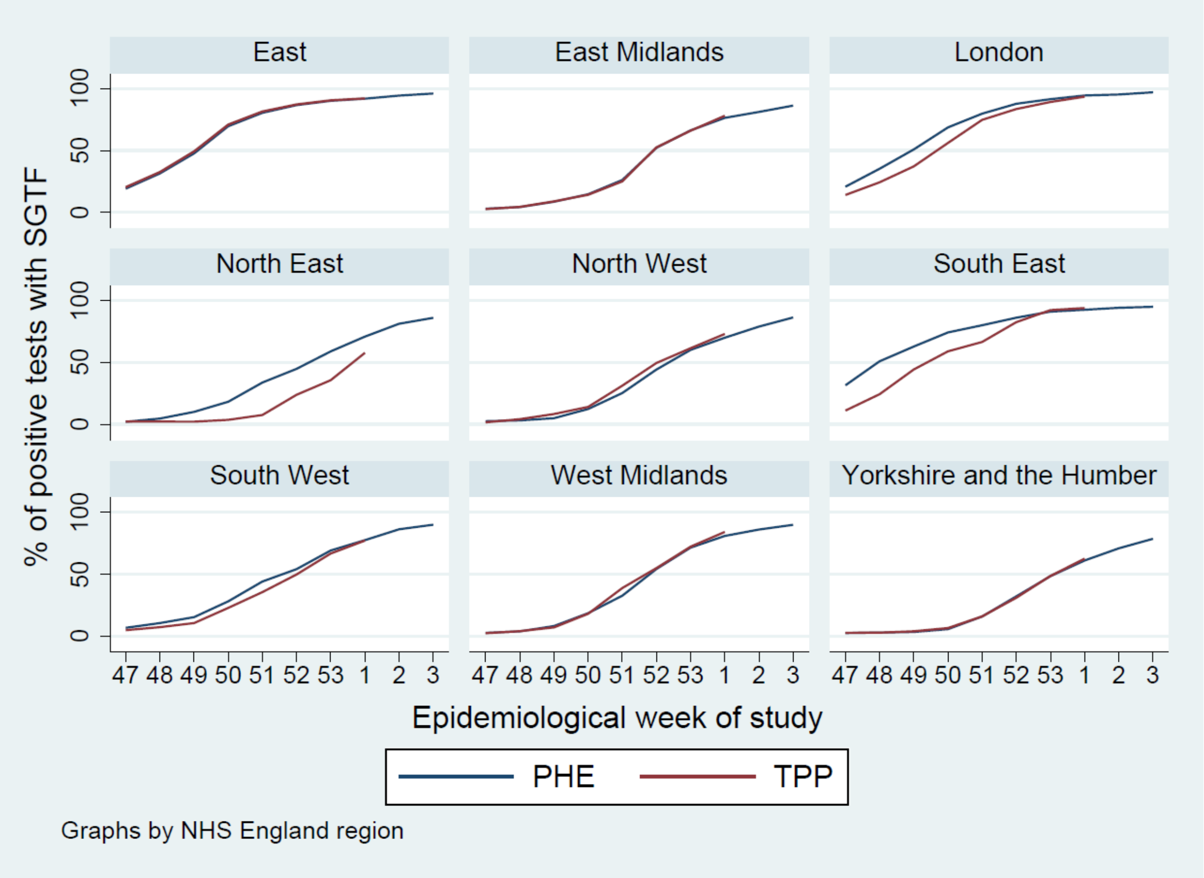


**Figure S1**. The percentage of positive tests in PHE reports[7] of SGTF compared with the proportion of tests in OpenSAFELY at the NHS region level.

#### 8. References

1. BETA -Data Security Standards - NHS Digital. NHS Digital. https://digital.nhs.uk/about-nhs-digital/our-work/nhs-digital-data-and-technology-standards/framework/beta---data-security-standards.
2. Data Security and Protection Toolkit - NHS Digital. NHS Digital. https://digital.nhs.uk/data-and-information/looking-after-information/data-security-and-information-governance/data-security-and-protection-toolkit.
3. ISB1523: Anonymisation Standard for Publishing Health and Social Care Data - NHS Digital. NHS Digital. https://digital.nhs.uk/data-and-information/information-standards/information-standards-and-data-collections-including-extractions/publications-and-notifications/standards-and-collections/isb1523-anonymisation-standard-for-publishing-health-and-social-care-data (accessed 26/2/2021.
4. Secretary of State for Health and Social Care - UK Government. Coronavirus (COVID-19): notification to organisations to share information. 2020. https://web.archive.org/web/20200421171727/https://www.gov.uk/government/publications/coronavirus-covid-19-notification-of-data-controllers-to-share-information (accessed 26/2/2021.
5. Department of Health and Social Care. Guidance: COVID-19 testing data: methodology note. 21/8/2020 2020. https://www.gov.uk/government/publications/coronavirus-covid-19-testing-data-methodology/covid-19-testing-data-methodology-note (accessed 25/2/2021.
6. Curtis HJ, MacKenna B, Croker R, et al. OpenSAFELY NHS Service Restoration Observatory 1: describing trends and variation in primary care clinical activity for 23.3 million patients in England during the first wave of COVID-19. *medRxiv* 2021: 2021.01.06.21249352.
7. Public Health England. Investigation of novel SARS-CoV-2 variant: Variant of Concern 202012/01. Technical briefing document on novel SARS-CoV-2 variant. https://www.gov.uk/government/publications/investigation-of-novel-sars-cov-2-variant-variant-of-concern-20201201 (accessed 25/2/2021.

### 9. Study Protocol

#### Case fatality risk of the SARS-CoV-2 variant of concern B.1.1.7 in England

**Version:** v1.0

**Date:** 22 Feb 2021

| Version history | Date | Comment |
|---|---|---|
| 0.1 | 1 Feb 2021 | Initial draft created |
| 0.2 | 8 Feb 2021 | Refinements following comments received |
| 0.3 | 11 Feb 2021 | Addition of causal framework and DAG section. Decided to drop absolute risk difference models in favour of relative risk models. Absolute risk to be calculated from predicted values from relative risk models or logistic regression if convergence issues. |
| 0.4 | 19 Feb 2021 | Extended study period by one week |
| 1.0 | 22 Feb 2021 | Added references and finalised |

### Contents

Background 12

Objectives 12

Study Design and Population 12

Inclusion criteria 13

Exclusion criteria 13

Causal framework 13

Study Measures 14

Exposure 14

Outcomes 14

Covariates 14

Statistical Methods 16

Baseline characteristics 16

28-day all-cause mortality 16

Cox proportional hazards regression 16

Covariate adjustment 16

Defining regions 17

*A priori* subgroup analyses 17

Sensitivity and additional analyses 18

Software and reproducibility 18

Feasibility and power calculations 18

Strengths and Limitations 18

Risk models fail to converge 18

Ethics 19

Conflicts of Interests 19

References 19

#### Background

The SARS-CoV-2 (COVID-19) variant of concern B.1.1.7 (VOC) was first identified in Kent, UK in autumn 2020. Early analysis suggests the VOC is more transmissible and it has since become the dominant strain throughout the UK. Only a small number of VOC cases are identified by whole-genome sequencing. Spike gene target failure (SGTF) has been adopted as a proxy for identifying VOC and has been shown to identify the VOC in more than 95% of cases during the period 16th November – 11th January.[1]

Studies using Public Health England (PHE) line listing data, hospital admissions and ONS death data have assessed the relative fatality of the VOC compared to the originally circulating viral strain (non-VOC) and have consistently demonstrated an increase in mortality associated with the VOC.[2,3] Although these studies were able to account for age, sex, ethnicity, deprivation index, time period and geographical area, they were unable to account for comorbidities, which have been shown to be strongly associated with death among those diagnosed with COVID-19.[4]

##### Objectives

To estimate the risk of death following confirmation of SARS-CoV-2 infection, comparing the risk among those infected with the VOC to those infected with the non-VOC accounting for both demographic factors and comorbidities. The risk of death will be quantified using the following methods:

i. A relative risk model, estimating relative and absolute 28-day all-cause mortality risk
ii. A Cox Proportional hazards regression model, estimating a hazard ratio

#### Study Design and Population

We will use a cohort study design nested within the OpenSAFELY platform. Using test result data from the PHE Second Generation Surveillance System (SGSS), we will select all those people who are:

(1) positive for SARS-CoV-2 based on PCR swab test results in the time window 16st November to 11th January and
(2) have data on SGTF

The study will focus on the comparison between those with SGTF and those without. Inconclusive SGTF results are considered in the sensitivity and additional analyses section.

The primary analysis will focus on a 28-day all-cause mortality outcome. All-cause mortality will be determined from ONS death data and is expected to have full ascertainment of deaths with a 2-week delay. Therefore, the 28-day all-cause mortality analysis will include all individuals with at least 42-days follow-up from the date of COVID-19 diagnosis to the date of last ONS death data upload (28-days plus 14-days to account for the delay in the ONS death data).

For Cox proportional hazards analysis all individuals will be included. Follow-up will be censored two weeks prior to the date of ONS death data upload for those without a documented date of death.

#### Inclusion criteria

- A positive SARS-CoV-2 PCR swab test result in SGSS within the window 16th November to 11th January
- Data available on SGTF in SGSS.
- Registered with a primary care practice using The Phoenix Partnership (TPP) software on the date of COVID-19 diagnosis, with at least one year of continuous GP registration.

#### Exclusion criteria

- Missing age, sex, or index of multiple deprivation, as these are indicators of poor data quality.
- COVID-19 diagnoses prior to the diagnosis in the study time window (based on either a positive test for SARS-CoV-2 in SGSS data or a diagnosis for COVID-19 in primary care).
- Receipt of vaccination against COVID-19 prior to diagnosis in the study time window.

#### Causal framework

The motivation for adjusting for demographics and comorbidities is not that they impact on the variant of COVID-19 infection *per se*, but are likely to be associated with the upstream process of getting a test (e.g. test-seeking behaviour, ability to access testing facilities). Therefore, adjustment attempts to correct for imbalances between the VOC and non-VOC exposure groups with respect to factors associated with getting a test. With the study population defined by SARS-CoV-2 positive test and SGTF data available, the minimum sufficient adjustment set implied by **Figure 1** is:

Age, care home status, comorbidities, deprivation index, smoking status.

**Figure 1 Causal framework DAG**

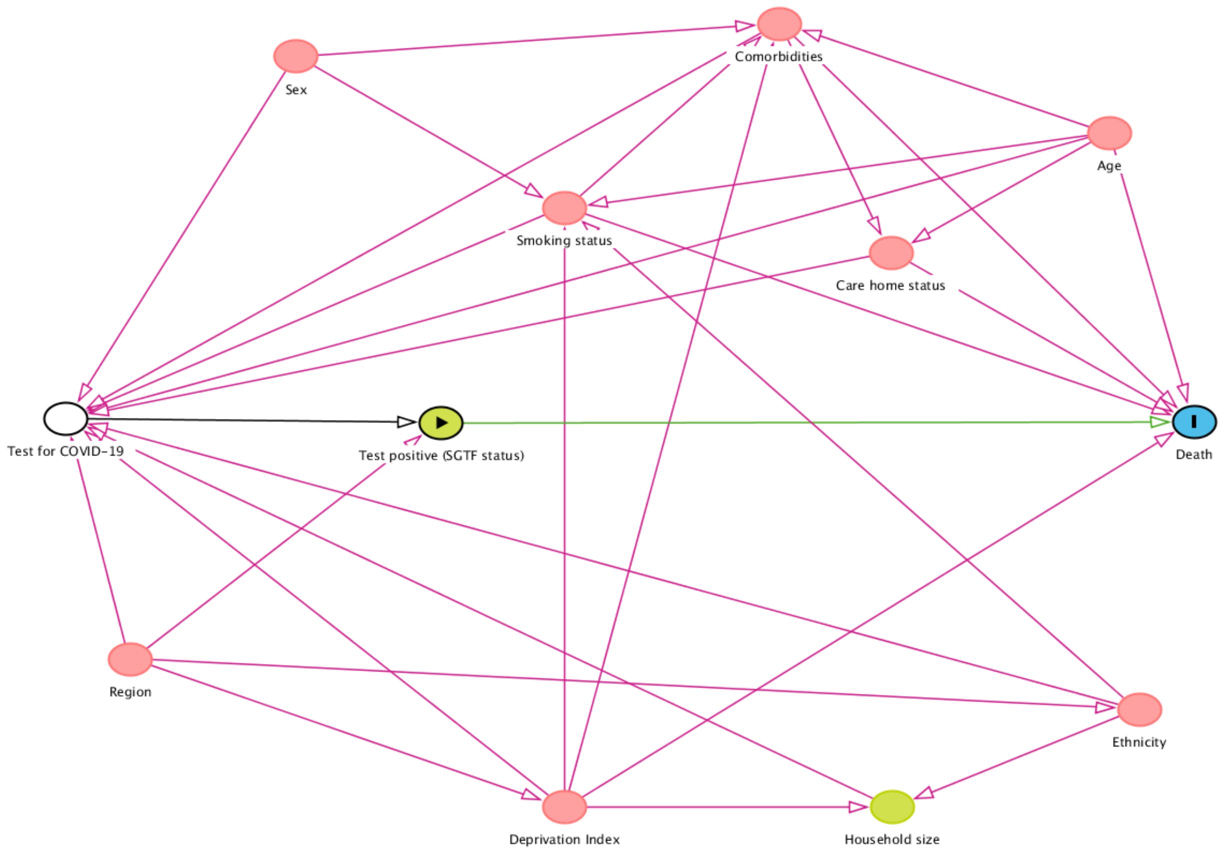

#### Study Measures

##### Exposure

SGTF on SARS-CoV-2 PCR swab test from SGSS data, referred to as the VOC exposure group. The comparator group being SARS-CoV-2 diagnoses without SGTF in SGSS data, referred to as the non-VOC group.

##### Outcomes

Death from any cause.

##### Covariates

Age, sex, deprivation index, ethnicity, smoking status, household size.

Region, defined by middle layer super output area (MSOA) from patient post code, or NHS England region.

Rural and urban location classification, and care home status.

Epidemiological week of the positive test.

Comorbidities: obesity, hypertension, chronic respiratory diseases other than asthma, chronic heart disease, diabetes, non-haematological and haematological cancer, reduced kidney function, chronic liver disease, stroke, dementia, other neurological disease, organ transplant, asplenia, rheumatoid arthritis/lupus, psoriasis, and other immunosuppressive conditions.

Table 1 **Study measures**

| Exposures | OpenCodelists Definition |
|---|---|
| SGTF | PHE Second Generation Surveillance System (SGSS) |
| Outcomes | |
| Death | All-cause registered deaths, from ONS |
| Covariates | |
| Ethnicity | https://codelists.opensafely.org/codelist/opensafely/ethnicity/ |
| Region | MSOA and STP are extracted from patient post code<br>UTLA regions are found using these look-up tables:<br>https://geoportal.statistics.gov.uk/datasets/lower-layer-super-output-area-2011-to-upper-tier-local-authorities-2019-lookup-in-england-and-wales-<br>https://geoportal.statistics.gov.uk/datasets/9f4c270148014f20bf24abff9a7aef62_0 |
| Comorbidities | https://codelists.opensafely.org/codelist/opensafely/aplastic-anaemia/<br>https://codelists.opensafely.org/codelist/opensafely/asplenia/<br>https://codelists.opensafely.org/codelist/opensafely/asthma-diagnosis/<br>https://codelists.opensafely.org/codelist/opensafely/asthma-inhaler-salbutamol-medication/2020-04-15/<br>https://codelists.opensafely.org/codelist/opensafely/asthma-inhaler-steroid-medication/2020-04-15/ |

| | |
|---|---|
| | https://codelists.opensafely.org/codelist/opensafely/asthma-oral-prednisolone-medication/2020-04-27/<br>https://codelists.opensafely.org/codelist/opensafely/bone-marrow-transplant/2020-04-15/<br>https://codelists.opensafely.org/codelist/opensafely/cancer-excluding-lung-and-haematological/2020-04-15/<br>https://codelists.opensafely.org/codelist/opensafely/chemotherapy-or-radiotherapy-updated/2020-04-15/<br>https://codelists.opensafely.org/codelist/opensafely/chronic-cardiac-disease/2020-04-08/<br>https://codelists.opensafely.org/codelist/opensafely/chronic-respiratory-disease/2020-04-10/<br>https://codelists.opensafely.org/codelist/opensafely/chronic-liver-disease/2020-06-02/<br>https://codelists.opensafely.org/codelist/opensafely/dementia/2020-04-22/<br>https://codelists.opensafely.org/codelist/opensafely/diabetes/2020-04-15/<br>https://codelists.opensafely.org/codelist/opensafely/chronic-kidney-disease/2020-04-14/<br>https://codelists.opensafely.org/codelist/opensafely/gi-bleed-or-ulcer/2020-04-08/<br>https://codelists.opensafely.org/codelist/opensafely/haematological-cancer/2020-04-15/<br>https://codelists.opensafely.org/codelist/opensafely/hiv/2020-07-13/<br>https://codelists.opensafely.org/codelist/opensafely/hypertension/2020-04-28/<br>https://codelists.opensafely.org/codelist/opensafely/inflammatory-bowel-disease/2020-04-07/<br>https://codelists.opensafely.org/codelist/opensafely/lung-cancer/2020-04-15/<br>https://codelists.opensafely.org/codelist/opensafely/other-neurological-conditions/2020-06-02/<br>https://codelists.opensafely.org/codelist/opensafely/permanent-immunosuppression/2020-06-02/<br>https://codelists.opensafely.org/codelist/opensafely/ra-sle-psoriasis/2020-04-14/<br>https://codelists.opensafely.org/codelist/opensafely/sickle-cell-disease/2020-04-14/<br>https://codelists.opensafely.org/codelist/opensafely/smoking-clear/2020-04-29/<br>https://codelists.opensafely.org/codelist/opensafely/smoking-unclear/2020-04-29/<br>https://codelists.opensafely.org/codelist/opensafely/solid-organ-transplantation/2020-04-10/<br>https://codelists.opensafely.org/codelist/opensafely/stroke-updated/2020-06-02/<br>https://codelists.opensafely.org/codelist/opensafely/temporary-immunosuppression/2020-04-24/ |

#### Statistical Methods

##### Baseline characteristics

Participant characteristics, including all covariates listed above, will be described at baseline (the date of positive SARS-CoV-2 test), comparing the two exposure groups (VOC and non-VOC). Continuous variables will be summarised by the mean and standard deviation and compared with a t-test, or median and interquartile range and Wilcoxon signed-rank test, as appropriate. Categorical variables will be summarized by the number and proportion in each group (n (%)) and compared with a chi-square test.

The median time-to-death and interquartile range of those who die will be presented by exposure.

The proportion of SARS-CoV-2 positive tests with SGTF, identifying the VOC, will be plotted over the study period by NHS England region and descriptively compared to PHE data for the whole population.

##### 28-day all-cause mortality

Case fatality risk will be calculated at 28-days post SARS-CoV-2 positive test result. Therefore, only those with 28-days of follow-up or a date of death within this window will be included in this analysis.

The relative risk for VOC cases vs. non-VOC will be calculated from a generalised linear regression model with binomial distribution and log link function. Absolute risk will be estimated by the predicted risk from this model.

**POST ANALYSIS NOTE:** there were model convergence issues when working on the risk scale. As per the limitations section, 28-day risk was therefore calculated from a logistic regression model.

##### Cox proportional hazards regression

The relative hazard of death for SGTF cases vs. non-SGTF will be calculated from a Cox proportional hazards regression model, with no requirement for 28-days follow-up time. Follow-up will be censored at the earliest of two weeks prior to the date of ONS death data upload or 7 days prior to receipt of a COVID-19 vaccination.

The hazard of death following a SARS-CoV-2 positive test result is expected to vary considerably between regions in England over time. Consequently, adjustment for region is unlikely to satisfy the proportional hazards assumption of a Cox model. To account for this variability, we will stratify the analysis on region, allowing a separate baseline hazard to be estimated for each region, but with covariate effects estimated over the full population – a stratified Cox PH model. The definition of regions is discussed below.

##### Covariate adjustment

Unadjusted, demographically adjusted, and fully adjusted estimates will be presented for each analysis.

Demographically adjusted models will include adjustment for the following covariates: Age will be included as a cubic spline term. Ethnicity will be grouped into five categories. The primary analysis will exclude patients with missing ethnicity. Sex, deprivation index, household size, and type of residence will be included as categorical terms.

Epidemiological week of the baseline SARS-CoV-2 positive test will be included as a categorical variable.

Region will be defined by UTLA, unless data sparsity prevents this level of granularity. In which case region will be defined by STP, or aggregated geographical areas defined by NHS England region. Rural or urban location classification will be included as a categorical variable with 5 levels in line with other work.

Fully adjusted models will additionally adjust for patient comorbidities, smoking status, and obesity status. Comorbidities will be aggregated into a categorical term taking values none, 1, and 2 or more. In line with previous work on the risk of death from COVID-19 on the OpenSAFELY platform. For smoking and obesity, missing values will be categorised as never smoked and no evidence of obesity, in line with previous OpenSAFELY studies.[4,5]

The causal framework indicates both these adjustment sets result in a causal estimate of the effect of the VOC on mortality. For comparison, we will also fit a model using the minimum sufficient adjustment set implied by the causal DAG (Age, comorbidities, deprivation index, smoking status).

#### Defining regions

Regional stratification will be a key consideration due to variability in the incidence of COVID-19 outcomes over time. Regions will be defined using patient middle super output area (MSOA) codes derived from patient post codes. MSOA data will be aggregated into upper tier local authority areas (UTLA) which will be the primary definition of regions for analysis.

Sensitivity analysis will define region by sustainability and transformation partnership (STP) areas, also defined from patient post codes. This analysis will assess the impact of regional definitions on the estimated risk and hazard of death for SGTF cases vs. non-SGTF.

Should data sparsity preclude regional adjustment at the UTLA and STP level, aggregated geographical areas defined by NHS England region will be used instead.

#### *A priori* subgroup analyses

Case fatality relative and absolute risk will be estimated in subgroups of *a priori* interest, after adjustment for confounding. Differences in risk in these subgroups will be formally tested with a likelihood ratio test for an interaction with SGTF exposure status.

The subgroups of interest to be assessed are:

- Age group (<50; 50-64; 65-74; 75-84; 85+)
- Ethnicity (in 5 categories)
- Comorbidity status (none, 1, 2+)
- Deprivation index (deciles of deprivation)
- Epidemiological week of positive SARS-CoV-2 test (each week of the study period, fortnightly if data are sparse)
- NHS England region (East, London, South East, South West, Midlands, North East and Yorkshire, North West)

#### Sensitivity and additional analyses

##### 40-day all-cause mortality

Differential time from positive SARS-CoV-2 test to death by SGTF exposure status has the potential to bias the analysis of risk. An additional analysis will consider an increase in the risk period to 40-days to assess the sensitivity of the findings to the risk period definition.

##### Inconclusive SGTF results

SGTF flags will be inconclusive in some cases. SGTF data are expected to take the values yes, no, maybe, unknown. The primary analysis will focus on the comparison of the yes group (VOC) with the no group (non-VOC). In additional analysis the risk of death in the maybe and unknown groups will also be quantified and compared to that of the VOC and non-VOC exposure groups.

##### Multiple imputation of missing ethnicity

Previous work in OpenSAFELY has identified that ethnicity data are missing for up to one quarter of all patients. The primary analysis will use the complete case set with regards to ethnicity. This sensitivity analysis will assess the impact of excluding records with missing ethnicity by imputing missing ethnicity using multiple imputation based on all variables included in the full adjustment set.

#### Software and reproducibility

Data management will be performed using Python and Google BigQuery, with analysis carried out using Stata 16.1 / Python. Code for data management and analysis as well as codelists archived online https://github.com/opensafely/sgtf-cfr-research.

#### Feasibility and power calculations

To be assessed when SGTF data are available.

### Strengths and Limitations

#### Risk models fail to converge

Although the size of our study population will likely be considerable with a large number of deaths, it's possible that the models of risk may fail to converge due to data sparsity in some covariate subsets. If this is the case we will revert to a logistic regression approach, estimating the log odds ratio. Inference on the risk of death will then be performed by converting predicted odds to estimates of absolute risk.

#### Non-random availability of SGTF data

Although the fact that we adjust for factors associated with getting tested should help account for possible non-random availability of SGTF data, we will also compare the characteristics of people included in the study (who all have SGTF data) with those not included in the study due to lack of SGTF data. This will help us assess whether those with SGTF are representative of all those tested during the time period of the study, and allow us to discuss the implications of this in our write-up as necessary.

#### Ethics

This study was approved by the Health Research Authority (REC reference 20/LO/0651) and by the LSHTM Ethics Board (reference 21863).

#### Conflicts of Interests

None to declare

#### References


1. Chand M. Investigation of novel SARS-COV-2 variant: Variant of Concern 202012/01 (PDF). *Public Health England PHE* 2020.
2. NERVTAG. NERVTAG paper on COVID-19 variant of concern B.1.1.7: Paper from the New and Emerging Respiratory Virus Threats Advisory Group (NERVTAG) on new coronavirus (COVID-19) variant B.1.1.7. https://www.gov.uk/government/publications/nervtag-paper-on-covid-19-variant-of-concern-b117.
3. Iacobucci G. Covid-19: New UK variant may be linked to increased death rate, early data indicate. British Medical Journal Publishing Group; 2021.
4. Williamson E, Walker AJ, Bhaskaran K, et al. OpenSAFELY: factors associated with COVID-19-related hospital death in the linked electronic health records of 17 million adult NHS patients. *MedRxiv* 2020.
5. Bhaskaran K, Bacon S, Evans S, et al. Factors associated with deaths due to COVID-19 versus other causes: population-based cohort analysis of UK primary care data and linked national death registrations within the OpenSAFELY platform. medRxiv 2021: 2021.01.15.21249756.
